# Phenotype Risk Scores: moving beyond ‘cases’ and ‘controls’ to classify psychiatric disease in hospital-based biobanks

**DOI:** 10.1101/2021.01.25.21249615

**Authors:** Dannielle S. Lebovitch, Jessica S. Johnson, Hillary R. Dueñas, Laura M. Huckins

## Abstract

Current phenotype classifiers for large biobanks with coupled electronic health records EHR and multi-omic data rely on ICD-10 codes for definition. However, ICD-10 codes are primarily designed for billing purposes, and may be insufficient for research. Nuanced phenotypes composed of a patients’ experience in the EHR will allow us to create precision psychiatry to predict disease risk, severity, and trajectories in EHR and clinical populations. Here, we create a phenotype risk score (PheRS) for major depressive disorder (MDD) using 2,086 cases and 31,000 individuals from Mount Sinai’s biobank BioMe ™. Rather than classifying individuals as ‘cases’ and ‘controls’, PheRS provide a whole-phenome estimate of each individual’s likelihood of having a given complex trait. These quantitative scores substantially increase power in EHR analyses and may identify individuals with likely ‘missing’ diagnoses (for example, those with large numbers of comorbid diagnoses and risk factors, but who lack explicit MDD diagnoses).

Our approach applied ten-fold cross validation and elastic net regression to select comorbid ICD-10 codes for inclusion in our PheRS. We identified 158 ICD-10 codes significantly associated with Moderate MDD (F33.1). Phenotype Risk Score were significantly higher among individuals with ICD-10 MDD diagnoses compared to the rest of the population (Kolgorov-Smirnov p<2.2e-16), and were significantly correlated with MDD polygenic risk scores (R^2^>0.182). Accurate classifiers are imperative for identification of genetic associations with psychiatric disease; therefore, moving forward research should focus on algorithms that can better encompass a patient’s phenome.

## Introduction

Substantial advances have been made in understanding the genetic architecture of psychiatric disease through the collection of very large case-control samples. Maximizing homogeneity within these samples (for example, applying stringent inclusion/exclusion criteria to define cases and controls), and minimizing variability in analytical design and diagnostic definitions, has led to the discovery of hundreds of genomic loci significantly associated with psychiatric disease. For example, the Psychiatric Genomics Consortium (PGC) has collated > 200,000 samples, across 11 psychiatric disorders, to identify 500 risk loci associated with these disorders^1^. The addition of biobank data to these analyses has substantially increased sample size, and identified yet more disease-associated variants; however, these findings have yet to yield meaningful insights into patient outcomes. To date, we lack the ability to stratify patients in clinically useful ways (for example, according to severity or chronicity), or to pinpoint appropriate treatments, or expected treatment outcomes, for each patient.

Arguably, as a field we must now enter the ‘translational’ stage of our research. Rather than exclusively focusing on assembling new, large cohorts, we must now consider the presentation of psychiatric disease within our clinics; and, by extension, within our medical records. Understanding the role of genetic associations within the clinical population may better elucidate the genetic architecture and biological mechanisms of disease; more importantly, identifying variants that are associated with disease outcomes or trajectories within our clinics may lead to more readily interpretable or actionable findings.

Extending our genetic analyses to incorporate EHR and biobank-based data will require careful consideration of phenotype definition. The binary concepts of ‘cases’ and ‘controls’ may become inappropriate when considering lifelong, common, episodic psychiatric traits, or traits that are characterized by a constellation of symptoms and behaviours. By contrast, hospital-based biobanks offer the opportunity to study the whole phenome over the lifespan of a patient^2–5^. Rather than pursuing single phenotype associations assessed at single time points, we can include trajectories of disease, comorbidities, and behaviours.

Here, we describe the creation of a ‘phenotype risk score’ for psychiatric traits, adapting the approach outlined by Bastarache et al^6^. Construction and use of phenotype risk scores (PheRS) may increase power in EHR studies, and provide whole-phenome perspectives of disease. Initially, this approach allowed symptom-based scoring of children with syndromic and mendelian traits in order to identify overlap and subtypes among disorders^6^. We extend this approach to psychiatric traits, and demonstrate application to major depressive disorder (MDD), a serious psychiatric disorder presenting with heterogeneous constellations of symptoms and comorbidities^7^. In particular, MDD is commonly comorbid with secondary psychiatric disorders (eg, substance use disorder^8^, generalized anxiety disorder^9^), as well as physical symptoms and traits (for example, cardiovascular disease^10^). In order to create our MDD-PheRS, we identify and quantify the comorbid traits and disorders commonly presenting with MDD; we term this the ‘comorbid phenome’, or *co-phenome*. MDD-PheRS scores are then calculated for each individual in our EHR as a weighted sum of co-phenome codes. Individuals with high scores have many of the traits and disorders that have been found to be commonly comorbid with MDD. Individuals with low scores display none, or almost none, of the traits and symptoms commonly comorbid with MDD.

We propose that our phenotype risk score may be particularly useful in instances where patients have restricted access to healthcare, or face financial or social barriers in seeking out or accessing mental health treatment^11^. Whereas assessing these individuals based purely on ICD diagnoses may misclassify them as ‘controls’; a whole-phenome perspective may instead reveal that these patients exhibit very high levels of comorbid traits, or even meet some diagnostic criteria of the trait in question. Genetically, and biologically, we might therefore expect these individuals to fall closer to a ‘case’ definition; wrongly assigning them as ‘controls’ would significantly reduce power in our studies^12^.

Whereas GWAS have historically focused on European populations, to the detriment of cross-ancestry insights and translatability^13–16^, hospital-based biobanks provide a more representative overview of the local population. EHR-based analyses therefore provide an opportunity to advance insights into disease aetiology across populations. In particular, our Mount Sinai BioMe ™ biobank participants^16–18^ are deliberately sampled to include equal proportions of patients with African American, European American, and Hispanic American ancestries. This provides us with a unique opportunity to ensure that our algorithms are valid, and useful, for our entire patient population, without discrimination by ancestry. Although we do not expect to find substantial biological differences in disease aetiology between groups, we do expect differences in experience, environmental exposures, and clinician diagnostic bias to affect diagnoses. Indeed, EHR- and epidemiological analyses identify multiple racial disparities in diagnosis, presentation, and treatment^19–22^ of MDD^19–29^.

## Methods

### Participants

Participants were included from Mount Sinai’s BioMe ™ biobank. BioMe ™ participants agree to the sharing of clinical electronic health record (EHR) and genotype data. The Charles Bronfman Institute for Personalized Medicine at Mount Sinai Medical Center (MSMC) BioMe ™ BioBank^18^ is an EMR-linked biorepository drawing from more than 70,000 inpatients and 800,000 outpatients annually. The MSMC serves the diverse local communities of upper Manhattan, including Central Harlem (86% African American), East Harlem (88% Hispanic/Latino), and the Upper East Side (88% European American) with broad health disparities across these groups. BioMe ™ enrolled over 50,000 participants between September 2007 and January 2020, with proportionally 21% African American, 34% Hispanic/Latino, and 31% European American individuals. The BioMe ™ population reflects community-level disease burden and health disparities with broad public health impact. Biobank operations are fully integrated in clinical care processes, with direct recruitment from clinical sites, waiting areas, and phlebotomy stations by dedicated Biobank recruiters, independent of clinical care providers, prior to or following a clinician standard of care visit. Recruitment currently occurs at over 30 clinical care sites.

Electronic health records for individuals in BioMe ™ include International Classification of disease (ICD) 9 and 10 codes, with an average of 31.8 ICD-10 diagnoses per individual, an average EHR-length of 2,942 days, and an average of 81 independent visits^30^. There are 2,086 individuals with an ICD-10 diagnosis of F33 MDD (**Table 1**), defined as “Major Depressive Disorder, recurrent”, of which the largest single subgroup is “MDD, recurrent, moderate” (ICD10 code F33.1, 1, N=156; **Table 1**).

### Identifying the MDD co-Phenome

The MDD co-phenome represents the typical diagnostic history of an individual with MDD in our healthcare system. In order to maximize power, we focused here on the most common MDD ICD-10 code (F33.1, “Moderate Depression”). To calculate the MDD co-phenome, we first removed rare ICD-10 codes (codes occurring in <5 individuals; 8,565 out of 15,495 codes removed). Next, we identified all codes significantly comorbid with MDD F33.1 using Fisher’s exact test.

### Creating a phenotype risk score (PheRS)

ICD-10 codes employ a hierarchical structure to describe processes, states, traits and disorders. Given the high degree of similarity between codes, the longitudinal nature of electronic health records, and the difficult, iterative process of arriving at psychiatric diagnoses, many ICD codes may be co-linear or redundant. Failure to correct for this overlap may result in significant skew in our phenotype risk score estimates.

In order to adjust for this potential redundancies and co-linearity between codes, we adopted a penalized regression approach (elastic net^31–33^). Elastic Net Regression (ENR) is a combination of two regularization regression methods, least absolute shrinkage and selection operator (LASSO) and Ridge Regression. LASSO has a shrinkage function to reduce collinearity and select parameters, while Ridge Regression maintains the equation from becoming underfitted.

We employed a ten-fold cross-validation approach to avoid overfitting, and used the “cvglmnet” function included in the glmnet 2.0-16 package in R version 3.5.3^31^.

We calculate our MDD-phenotype risk score (MDD-PheRS) as a weighted sum of MDD-co-phenome ICD codes using two methods:

1. PheRSm1: Weights are calculated as the log-inverse prevalence of each associated ICD-10 diagnosis code with MDD in BioMe™ (**Box 1**), analogous to Bastarache et al^6^.
2. PheRSm2: Weights are defined as effect sizes (betas) obtained from the ten-fold cross validation elastic net model (**Box 1**).

### Box 1: PheRS Calculation

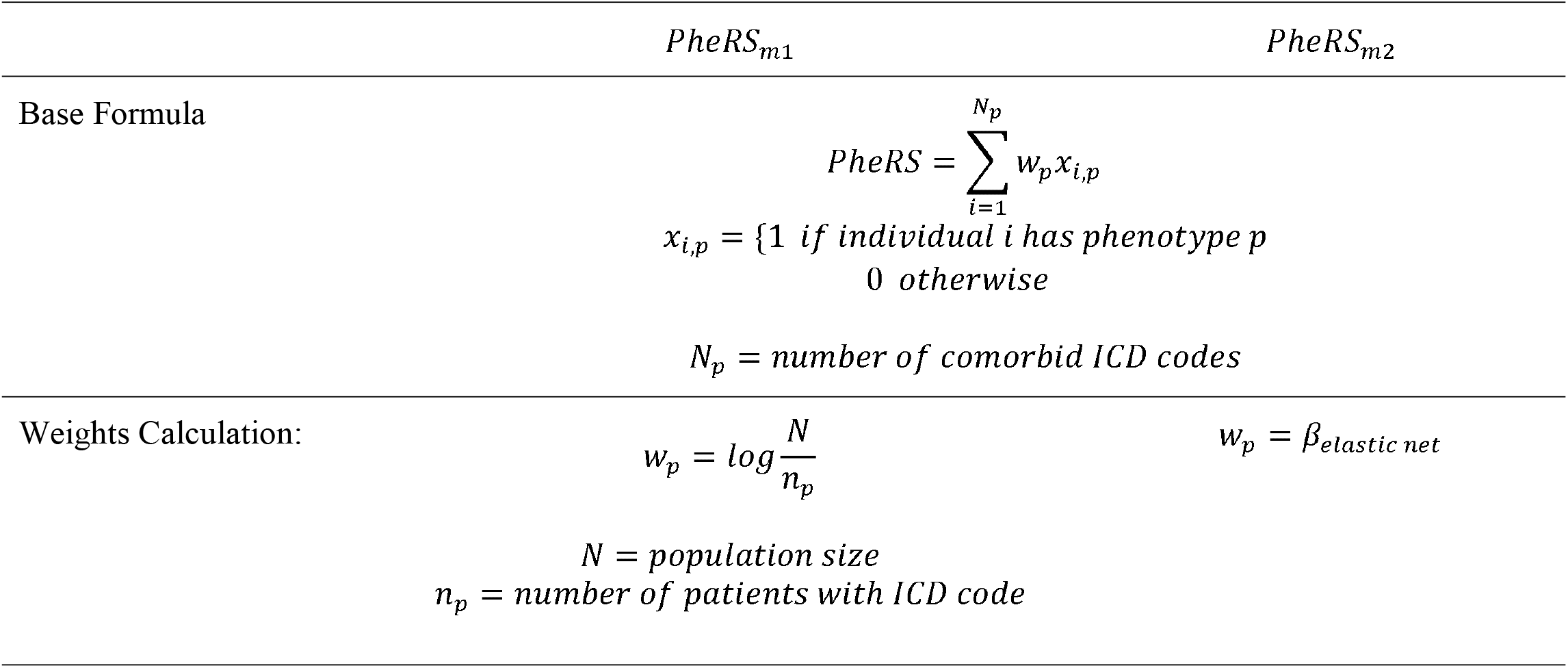

We compared PheRS distributions between (i) individuals with- and without-MDD F33.1 codes and (ii) individuals with- and without-**any** MDD diagnostic codes using kolgorov-smirnov tests for significance.

### Comparison to MDD Polygenic Risk Scores

We calculated MDD-polygenic risk scores (MDD-PRS) for all individuals in our biobank, using the most recent PGC summary statistics^34^ (142,000+ individuals; **45**,**396 cases and 97**,**250 controls**). PGC cases were identified using self-report, clinician interview and/or medical record diagnoses. We used PRSice-2^35,36^ to calculate MDD-PRS for all BioMe ™ participants at ten standard p-value thresholds: 1×10^−4^, 1×10^−3^, 0.01, 0.05, 0.1, 0.2, 0.3, 0.4, 0.5, 1.

We partitioned individuals into MDD-PRS deciles, and tested for case enrichment in the top decile compared to (i) the bottom decile and (ii) all other deciles. In particular, we tested for enrichment in the top decile of (1) MDD cases/control status as defined by ICD code F33.1; and fold change of (2) PheRSm1and (3) PheRSm2.

PRS analyses are particularly vulnerable to ancestry differences between ‘base’ and ‘target’ GWAS^14,37–40^; i.e., the ancestry of individuals from which the PRS is derived must closely match that to which the PRS is applied. To date, no large-scale MDD-GWAS have been performed in individuals of African-American or Hispanic descent. Therefore, we restricted this section of our analysis to individuals of European descent.

### Cross-ancestry specificity of phenotype risk scores

Our MDD co-phenome assesses the whole phenome of each individual with MDD in our healthcare system. These may reflect biological similarities or comorbidities between disorders, pleiotropy of genetic risk factors, diagnostic bias or mis-diagnosis, or simply patterns of healthcare usage that reflect, but are not directly related to, an underlying health condition^11^.

Modeling the whole EHR without accounting for potential differences between ancestries risks masking crucial differences that may affect the score, and ultimately may produce a less useful algorithm for understanding disease risk and clinical outcomes.

In short, we expect that a person’s race likely significantly shapes their access to and experience of healthcare systems^11^. Therefore, we repeated our co-phenome analysis, partitioning individuals by self-defined race/ethnicity. We included racial and ethnic groups with >1,000 individuals, and >100 MDD-F33.1 diagnosis counts (**S. Table 1;** categories with sufficient data for analysis are shown in **bold**).

For each group with sufficient individuals and cases, we repeated our co-phenome analysis using the same elastic net regression approach. In instances where co-phenomes differed significantly, we repeated construction of our MDD-PheRS. We compared distributions of MDD-PheRS using overall- and group-specific-co-phenome input and assessed significance using Kolgorov-smirnov tests.

Next, we constructed three additional PheRS, specific to each large ancestry group, and compared predictive accuracy within and across ancestries, and to an approach grouping across all ancestries (ie, our original PheRS). We first derived PheRS effect sizes using a ten-fold cross-validation approach in each ancestry group separately (the “base” population). Next, we applied these PheRS, and our overall PheRS, to each of the three ancestry groups in turn (the “target” populations), and tested F33.1 prediction accuracy across all base-target combinations.

## Results

### Characterizing the MDD co-phenome

We define the ‘co-phenome’ as the set of diagnoses and traits that are comorbid with depression within our healthcare system. First, we identified all depression-related codes in our biobank (**Table 1**). To maximize power, we selected the most frequent code (F33.1, Moderate Depression) to represent ‘depression’ within our EHR. We identified 158 ICD-10 codes that were significantly comorbid with F33.1-MDD (**Table 2**), including many well-studied MDD risk factors (e.g., childhood sexual abuse^41^, homelessness^42^), comorbidities (e.g., BMI^43^, respiratory distress^44^) and clinical features (e.g., suicidal ideation^45^, opioid abuse^46^), as well as some less expected associations (e.g., guillain-barre syndrome, rheumatic heart failure).

### Creation of an MDD-Phenotype Risk Score

Phenotype risk scores (PheRS) quantify the amount of MDD co-phenome codes present in each individuals’ diagnostic history. Higher PheRS scores imply higher whole-phenome similarity to MDD cases. In order to account for the significant diagnostic and biological overlap between ICD codes, the hierarchical, clustered nature of ICD-10 codes, the imprecision inherent in ICD-10 code usage, the longitudinal nature of EHR, and the difficult, iterative process of arriving at psychiatric diagnoses, we applied a penalized regression framework (elastic net regression) to automatically select codes for inclusion in our PheRS (**Figure 1**) using a ten-fold cross-validation framework.

**Figure 1:**
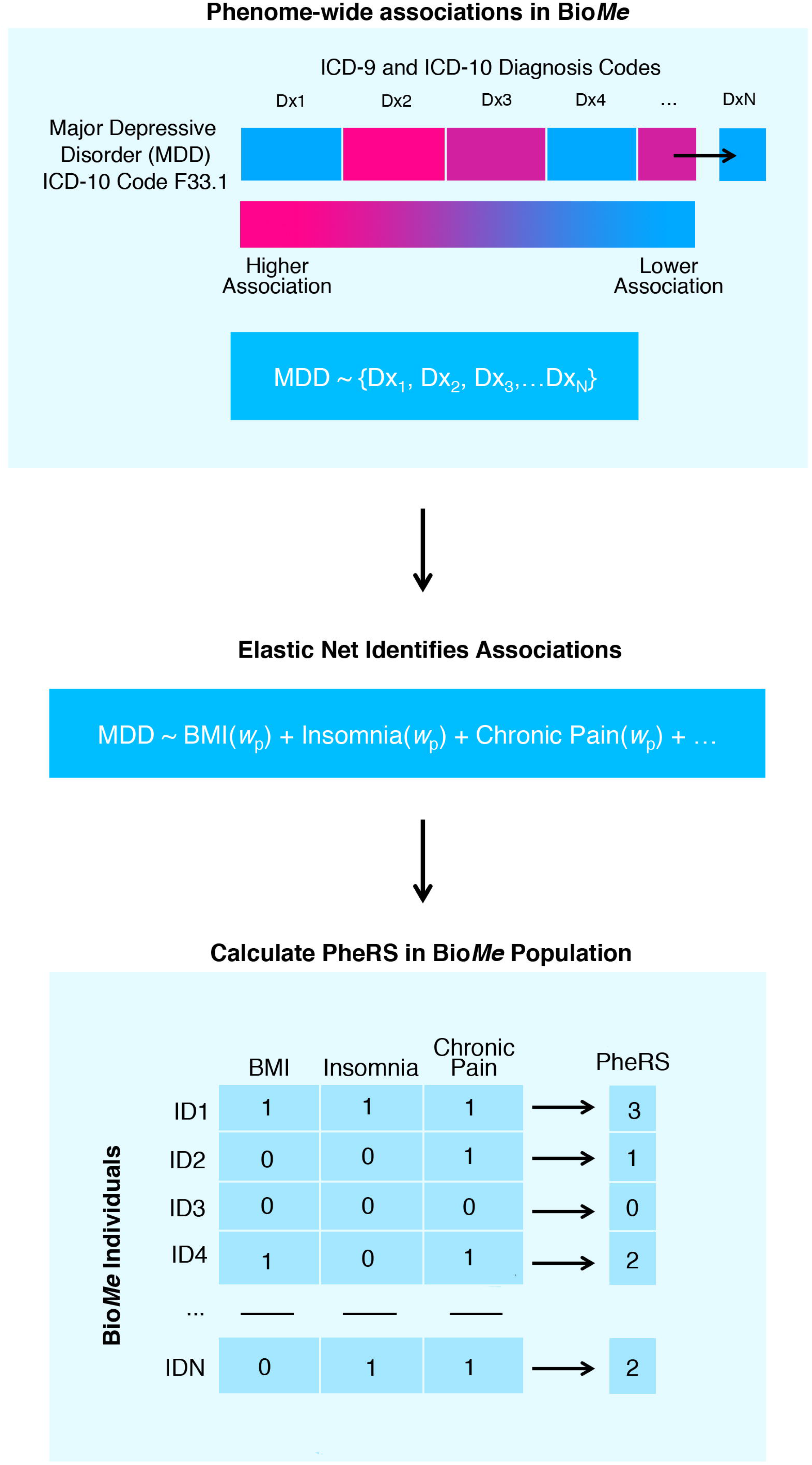
Constructing Phenotype Risk Scores from EHR data for complex traits. Step 1: Identification of traits, disorders and phenotypes associated with MDD. Step 2: Construction of a joint model predicting MDD from all comorbid traits, using 10-fold cross-validation and elastic net regression. Step 3: Phenotype risk scores are calculated as weighted sums of traits and disorders present in each individual’s phenome. Traits are weighted according to (i) prevalence (PheRSm1) or (ii) elastic net calculated effect size (PheRSm2).

Our model includes 158 ICD codes (**Table 3**), and significantly predicts F33.1 MDD in a ten-fold cross-validation framework (R^2^ = 0.367). The largest contributing effect size of all 158 codes identified is respiratory failure (J96.91, OR= 7.41), followed by personal history of physical or sexual abuse in childhood (Z62.810, OR= 6.03). Of the 158 codes identified to be predictive of F33.1 MDD, 30 are related to psychiatric disorders and traits; of these, the largest effect size is from PTSD (ICD-10 code F3.10, OR= 4.40). PheRS scores for individuals with (i) F33.1-MDD and (ii) broadly defined MDD (F32, F33) are significantly higher than those for patients without MDD diagnoses (kolgorov-smirnov test, p<2.2×10^−16^; **Figure 2**).

**Figure 2:**
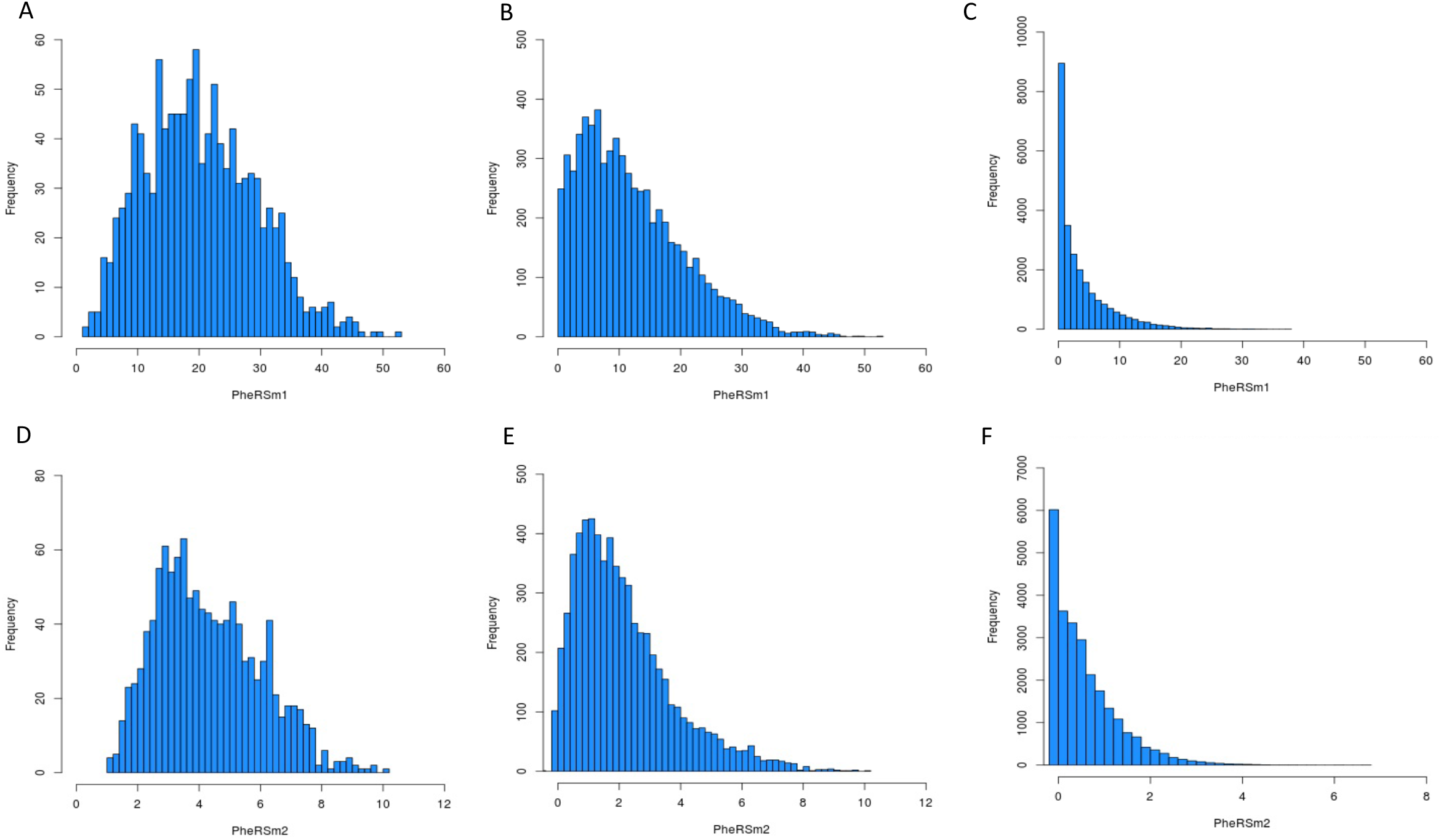
MDD-Phenotype Risk Scores are significantly higher among individuals with histories of depression. For both methods (PheRSm1-A-C; PheRSm2-D-F), we find that phenotype risk scores are highest among individuals with diagnoses of F33.1 (A, D); elevated among individuals with any depression diagnosis, excluding F33.1 (B, E), and lowest among individuals with no history of any depression diagnoses (C,F). All comparisons are significant; kolgorov-smirnov test p<2.2×10^−16^.

### MDD-PheRS correlate with polygenic risk scores

In order to test whether individuals with higher MDD phenotype risk scores also have higher genetic risk for MDD, we calculated MDD-PRS^34^ for all 10,792 European BioMe ™ individuals^18^ using PRSice-2^35,36^. Individuals in the top decile of MDD-PRS had significantly higher MDD-PheRS scores compared to individuals in the bottom decile (fold change=3.57, 2.46 for PheRSm1, PheRSm2, respectively; **Figure 3**). This enrichment persists even when removing individuals with any MDD diagnostic codes (ie, F32, F33) from our analysis (fold change=3.58, 3.21, respectively; **Figure 3**). Next, we compared the amount of phenotypic variance explained by MDD-PRS for both our phenotype risk scores, and MDD F33.1 case-control status. We find substantially more variance explained by MDD-PRS for both phenotype risk scores (R^2^=0.217, 0.182, respectively) than for F33.1 case-control status (R^2^=0.051).

**Figure 3:**
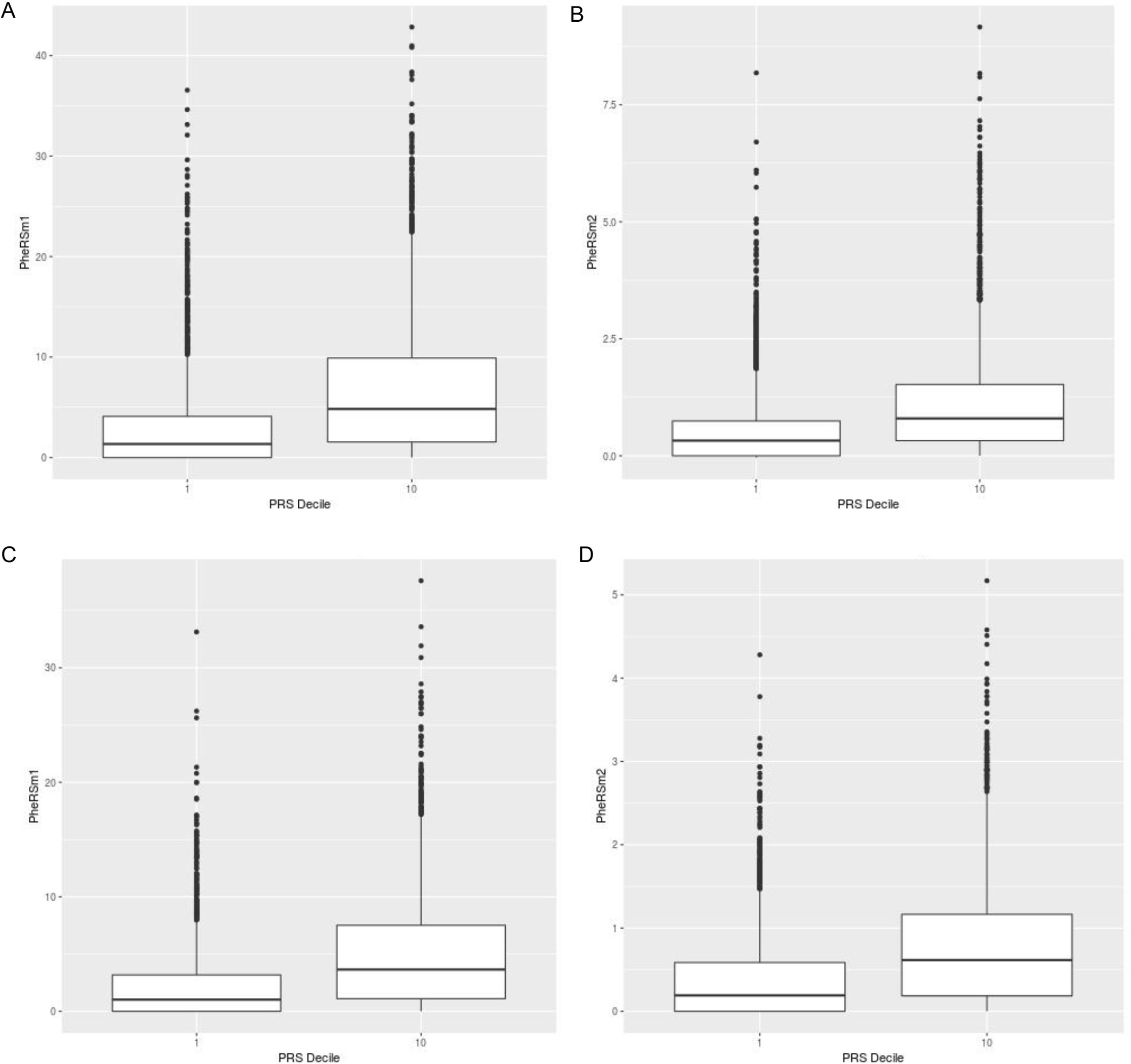
MDD Polygenic Risk Scores predict higher MDD Phenotype Risk Scores. Phenotype Risk Scores are significantly higher among individuals with the top decile of polygenic risk, compared to the bottom, for both methods (PheRSm1-**A**; PheRSm2-**B**). This effect persists even when removing individuals with F33.1 diagnoses (PheRSm1-**C**. Fold change =3.58; PheRSm2-**D**. Fold change=3.21).

### PheRS vary across ancestries

Although we do not expect substantial biological aetiological differences between racial/ethnic groups, it is likely that race and ethnicity plays a significant role in shaping our experiences of the healthcare system^11^. As such, we hypothesized that co-phenome profiles may vary when partitioning patients according to race. In **Figure 4i-iii**, we considered three potential scenarios. First-if bias plays only a small role in diagnoses, i.e, if the majority of diagnoses are assigned accurately, we expect highly overlapping comorbidity profiles and risk-factors for MDD between patient groups (**Figure 4i**). However, if bias affects diagnostic accuracy in one group compared to the others, we might expect less overlap in co-phenomes between groups (**Figure 4ii**); alternatively, bias may lead to differentially missing data between groups (for example, bias may lead to fewer lab tests or follow-up appointments; **Figure 4iii**).

**Figure 4:**
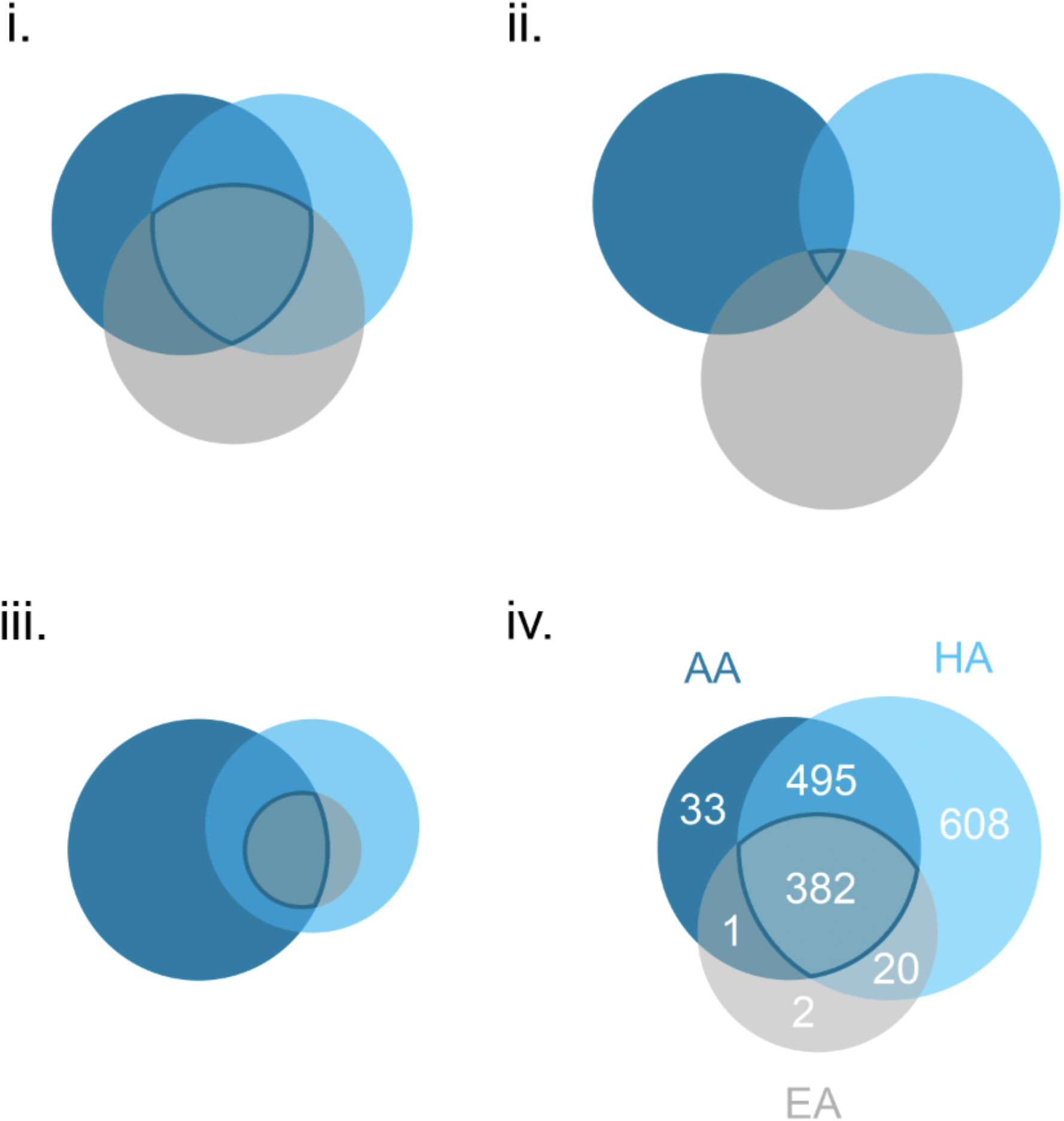
Hypothetical scenarios modelling the impact of diagnostic bias, compared to observed MDD-comorbidity profiles. (i) Bias plays only a small role in diagnoses, and the majority of diagnoses are assigned accurately. We expect highly overlapping co-phenomes and risk-factors for MDD between patient groups. (ii) Diagnostic bias affects these groups unevenly; we expect less overlap in co-phenomes. (iii) Diagnostic bias results in under-diagnosis and missing data (ie, a smaller than expected co-phenome) in one group, and uneven overlap between groups. (iv) Observed overlap between MDD co-phenomes of African-American (AA), European-American (EA), Hispanic-American (HA) in our database.

To test these hypotheses, we partitioned patients into three groups, according to self-reported race/ethnicity: European (N=10,792), African-American (N=8,046), Hispanic-American (N=11,047). We identified significantly different co-phenome profiles specifically among Hispanic-American patients within our system (**S. Table 2**). In particular, Hispanic-American individuals with MDD had very large numbers of co-morbid ICD codes, many of which (608) were only identified among this population. In addition, 495 codes were significant among African-American and Hispanic-American individuals with MDD, but were not co-morbid with MDD among European-Americans. By contrast, almost all comorbid codes among European-Americans were shared with both other groups (**Figure 4iv**).

Next, we constructed three additional PheRS, specific to each large ancestry group, and compared predictive accuracy of PheRS within and across ancestries (ie, our original PheRS), resulting in nine PheRS scores. For each score, the ‘base’ dataset describes the ancestry group used to derive the score, and the ‘target’ describes the dataset to which the score is applied; e.g., PheRS_AA-HA_ denotes a score developed in African-American patients and applied in Hispanic-American patients. We found significant correlation in PheRS scores across all base-target combinations (spearman rho > 0.58), with highest correlations for AA/HA base/target combinations (spearman rho > 0.79). We did not observe any effect of base population on predictive accuracy when applied to the full cohort (R^2^ range 0.196-0.211; **S. Table 3**); however, we do observe differences when applying across specific groups (eg: base-EA target-AA). As expected, we observe the highest predictive accuracy when base and target populations are matched (**S. Table 3**); followed by AA-HA base/target combinations. Scores derived in solely EA individuals performed noticeably worse in AA/HA populations, and vice-versa.

## Discussion

Large-scale, collaborative approaches to amass large numbers of cases and controls have yielded great insights into complex trait architecture. In particular, concerted efforts across the psychiatric genetics field has resulted in >500 genome-wide significant associated loci across 15 psychiatric disorders and traits. However, such case/control definitions match neither the lived experiences of patients, nor of clinicians. In particular, these study designs cannot account for comorbidities or whole phenome perspectives, nor account for changing comorbidity patterns and diagnostic journeys throughout the lifespan.

Electronic Health Records (EHR) provide an attractive opportunity to design studies that more closely match the experiences of our patients. In this study, we demonstrate application of phenotype risk scores to quantify EHR-MDD comorbidities. We suggest that these PheRS may be applied for two key goals. First, to increase statistical power in EHR analyses by assigning a quantitative ‘MDD-PheRS’ value to all individuals, rather than a binary case/control designation. Second, our analysis may allow us to identify individuals with ‘missing’ diagnoses; for example, those who exhibit a large number of comorbid diagnoses or risk factors, but who lack explicit MDD diagnoses within their records. Such missing diagnoses may stem from a lack of ability to afford treatment; an unwillingness to seek treatment due to stigma or shame discussing mental health^47^; diagnostic oversight or bias^48^; or difficulty in reaching an accurate diagnosis due to the lack of reliable tests or biomarkers for psychiatric disease^49^. Here, we suggest that such individuals may be identified using PheRS; for example we identified people with high PheRS and high MDD PRS but never received a formal diagnoses.

As we might expect for a complex, episodic trait, we find a wide range of associated traits and diagnoses contributing to our score. These associations can be grouped into a few broad categories. First, known biological risk factors for MDD, for example; Type II Diabetes (E11.65)^50^. Second, we identify a number of environmental factors (e.g., homelessness^51^), which may precipitate or occur as a consequence of MDD. Third, we identified a number of psychiatric diagnoses and traits (e.g. Alcohol Dependence; F10.21). These may represent truly distinct diagnoses (for example, patients may truly have distinct PTSD and MDD diagnoses); or, they may provide evidence of complex diagnostic journeys for patients within our study. For example, early diagnoses of MDD may later be amended to diagnoses of bipolar disorder as more diagnostic information becomes available to clinicians. More complex evaluations with larger psychiatric EHR will be required to disentangle these patterns.

Finally, we identified a set of unexpected associations (moderate persistent asthma, J45.40; Gastro-esophageal reflux disease, K21.9). These may be representative of many factors. It is possible that some unknown or understudied shared biological aetiology exists between these traits; (for example, between lower leg pain (M79.604) and diabetes or homelessness, both of which are comorbid with MDD in our sample). It is also possible that some unknown ‘healthcare pleiotropy’ exists between traits; that is, traits that commonly co-present within the EHR for factors unrelated to biology-for example, sets of tests commonly performed jointly at annual check-ups.

Our analysis identifies a substantial number of individuals with high MDD PheRS and PRS values, but with no MDD diagnoses. Similarly, we identified some individuals with very low PheRS and PRS scores, but with multiple ICD-MDD diagnoses. Traditional EHR approaches, which typically require 1-2 diagnostic codes in order to form a diagnosis, would misclassify these individuals. For example, among individuals falling in the top two PheRS and PRS deciles, we identified 383 individuals with no F33.1 MDD diagnoses.

In order to ascertain whether our PheRS captures biologically relevant information about MDD, we tested whether PheRS scores within our biobank also correlated with MDD-PRS derived from the latest PGC-MDD GWAS. We found that our PheRS scores correlate significantly with MDD-PRS (Figure. 5), and explained more variance in the MDD-PheRS than in the directly observed MDD diagnosis. Notably, excluding individuals with MDD from our MDD-PRS analysis does not remove the association between PRS and PheRS, implying that this effect is not driven by individuals with MDD and high PheRS. Therefore, we hypothesize that either (i) our PheRS captures undiagnosed MDD cases, driving the PRS-PheRS associations; or (ii) that MDD-PRS is associated with the constellation of symptoms and traits associated with MDD (i.e., the MDD co-phenome), even in the absence of MDD.

Finally, we tested cross-ancestry applicability of our PheRS across three key racial/ethnic groups in our biobank. We urge caution in interpreting these results. Since race is a social rather than biological concept, we do not expect that true biological differences exist between self-defined racial groups; nor do we expect the genetic aetiology of MDD to differ according to race. However, we do expect that access to, and experience of, healthcare will differ according to race. In large part, these differences will stem from structural inequality, bias and racism experienced by our patients-in differing access to expert clinicians and insurance; in biased diagnostic practices and therapeutic approaches; and in differing environmental exposures and stressors. While some of these factors may be quantified within our medical records (for example, the number of visits, referrals to specialists or life-threatening traumas experienced), others will be implicit and unquantifiable (for example, biased or stereotyped diagnostic guidelines, or long-term stress from racist microaggressions). Sample sizes are insufficient here to probe effects of intersectionality (eg, compound bias experienced by Black women), although we expect these compound factors, as well as additional unknown biases inherent across healthcare systems, to further bias comorbidity profiles and diagnostic accuracy within EHR.

Indeed, we found significantly different comorbidity profiles between all three racial/ethnic groups included in our analysis; in particular, we found far more ICD codes co-morbid with MDD among Hispanic-Americans overall, and a large number of codes comorbid only among Hispanic-Americans. By contrast, almost no codes were comorbid uniquely among European-Americans: rather, all codes significantly co-morbid among European-Americans were also significantly comorbid among other groups. These differences imply that MDD diagnoses are applied differently across racial and ethnic groups. It is possible that these differences stem from power imbalances- that is, the increased number of MDD cases among Hispanic-Americans enables us to identify significantly more associations; however, this would not account for the large number of codes shared only between African-American and Hispanic-American individuals. It is also possible that the diagnosis in question (F33.1) is given more readily-that is, at some lower clinical threshold-among HA patients than in other groups. This broader, more general use of the diagnosis would also account for the larger and more diffuse set of comorbidities, and is in line with our previous research regarding diagnostic bias in psychiatry^11^.

We tested which approach is most appropriate using a cross-ancestry base/target design. We derived PheRS using (i) all samples; (ii) each of the three racial/ethnic groups only, and tested predictive accuracy (R^2^) in the full cohort, and in each ancestry separately. We observe highest accuracy when base and target populations are matched (as would be expected), and only slight reductions in accuracy with HA/AA base/target combinations. Importantly, these patterns do not match expectations based on sample size. By default, larger discovery (base) populations should result in more accurate phenotype risk scores. However, AA- and HA-base PheRS perform similarly, despite significant differences in sample size; and both outperform EA-base PheRS when applied to the full cohort. Our results indicate that it is not appropriate to assume that a European-default derived algorithm would be sufficient or appropriate.

However, two scenarios may justify construction of a PheRS using all samples jointly, regardless of race/ethnicity. First, it is possible that the power gained from increasing sample size (ie, by jointly analyzing all samples) outweighs the loss of specificity. However, it will be vital to ensure that power is gained evenly across all individuals and groups-ie, that predictive accuracy is not just increased significantly among a subset of participants. Alternatively, it is possible that diagnostic accuracy is higher among some racial/ethnic groups than others-that is, the MDD diagnosis underlying our model is more reliably applied in one group. In this case, PheRS derived from all samples, or from this single group, may perform best. More research into racial and ethnic biases in psychiatric diagnoses is warranted to probe these questions further. In particular, we caution that MDD diagnoses may be substantially less biased across ancestries than other psychiatric diagnoses. For example, we have previously shown that psychotic diagnosis and treatment are significantly different between racial and ethnic groups in our sample; similarly, previous studies have shown that diagnostic patterns differ greatly for eating disorders, autism spectrum disorder, and ADHD, among others. For these traits, race and ethnicity may be even more relevant in creating a useful phenotype risk score.

In summary, shallow phenotypes composed of a single data point from EHR do not accurately capture a patient’s experience with disease. Algorithms comprising multiple data points, such as PheRS, allow us to consider heterogeneous patient experience, comorbidity, bias, and ancestry, as well as a patient’s whole phenome, in order to arrive at accurate and robust phenotypes. With this tool, we begin to use our currently available biobanks to look for translational opportunities to better predict trajectories of disease, comorbidities, and behaviours to find multi-omic associations to understand disease etiology.

## Data Availability

Summary data is made available in the supplementary tables of this article.

## Acknowledgements

This work was supported by a Faculty Scholar Award from the Seaver Foundation: “Analytical Genomics of Vulnerable Populations”.

This work was supported in part through the resources and staff expertise provided by the Charles Bronfman Institute for Personalized Medicine and The BioMe ™ Biobank Program at the Icahn School of Medicine at Mount Sinai.

We thank Dr. Eli Stahl, and Dr. Alex Charney for their thoughtful comments during lab meetings and poster sessions. We thank Dr. Inga Peter for her helpful comments and revisions to this manuscript, and for her assistance in examining Ms. Lebovitch’s MPH thesis, which formed a preliminary version of this analysis.

We also acknowledge the supportive atmosphere of the Pamela Sklar Division of Psychiatric Genetics; our work is stronger for the collegiate and collaborative atmosphere of our division. As always, we are grateful for the advice, guidance and mentorship of our founder and namesake, Dr. Pamela Sklar.

## Table legends

Table 1: Number of BioMe Participants with ICD10 MDD diagnostic codes Table 2: ICD Codes significantly associated with F33.1

Table 3: ICD codes contributing to MDD-PheRS Supplementary table legends:

S. Table 1: Number of F33.1 Diagnoses according to self-reported sex, and race/ethnicity

S. Table 2A: ICD codes significantly co-morbid with MDD among African American patients

S. Table 2B: ICD codes significantly co-morbid with MDD among European American patients

S. Table 2C: ICD codes significantly co-morbid with MDD among Hispanic American patients Supplementary Table 3: Cross-Ancestry predictive accuracy

## Notes

### Competing Interest Statement

The authors have declared no competing interest.

### Author Declarations

Founded in September 2007, BioMe is a biobank that links genetic and electronic medical record (EMR) data for over 30,000 individuals recruited primarily in ambulatory care settings in the Mount Sinai Health System (MSHS) in New York City. The current study was approved by the Icahn School of Medicine at Mount Sinai's Institutional Review Board (Institutional Review Board 07-0529). All study participants provided written informed consent.

