## Supplementary material for "Phenotype Risk Scores: moving beyond ‘cases’ and ‘controls’ to classify psychiatric disease in hospital-based biobanks": Table 1

**Table 1: Number of BioMe Participants with ICD10 MDD diagnostic codes**

| **ICD Code** | **Diagnosis** | **Number of unique participants** |
| --- | --- | --- |
| **F33.0** | Major depressive disorder, recurrent, mild | 514 |
| **F33.1** | Major depressive disorder, recurrent, moderate | 1156 |
| **F33.2** | Major depressive disorder, recurrent severe without psychotic features | 399 |
| **F33.3** | Major depressive disorder, recurrent, severe with psychotic symptoms | 240 |
| **F33.4** | Major depressive disorder, recurrent, in remission | 663 |
| **F33.40** | …… unspecified | 66 |
| **F33.41** | Major depressive disorder, recurrent, in partial remission | 361 |
| **F33.42** | Major depressive disorder, recurrent, in full remission | 236 |
| **F33.8** | Other recurrent depressive disorders | 14 |
| **F33.9** | Major depressive disorder, recurrent, unspecified | 782 |
| **Total** |  | 3768 (2086) |
